# Accuracy of Rapid Point-of-Care Antibody Test in patients with suspected or confirmed COVID-19

**DOI:** 10.1101/2020.11.17.20233296

**Authors:** Rama Vancheeswaran, Merlin L Willcox, Beth Stuart, Matthew Knight, Hala Kandil, Andrew Barlow, Mayon Haresh Patel, Jade Stockham, Aisling O’Neill, Tristan W Clark, Tom Wilkinson, Paul Little, Nick Francis, Gareth Griffiths, Michael Moore

**Author notes:** Correspondence to: Dr Merlin Willcox, School of Primary Care, Population Sciences and Medical Education, University of Southampton Aldemoor Health Centre, Aldermoor Close, Southampton SO16 5ST.

## Abstract

**Objectives:** To assess the real-world diagnostic accuracy of the Livzon point-of-care rapid test for antibodies to SARS-COV-2

**Design:** Prospective cohort study

**Setting:** District general hospital in England

**Participants:** 173 Patients and 224 hospital staff with a history of COVID-19 symptoms, and who underwent PCR and/or reference antibody testing for COVID-19.

**Interventions:** The Livzon point-of-care (POC) lateral flow immunoassay rapid antibody test (IgM and IgG) was conducted at least 7 days after onset of symptoms and compared to the composite reference standard of PCR for SARS-COV-2 plus reference laboratory testing for antibodies to SARS-COV-2. The SARS-CoV-2 RT-PCR was tested using the available molecular technology during the study time (PHE laboratories, GeneXpert^®^ system Xpert, Xpress SARS-CoV-2 and Source bioscience laboratory). All molecular platforms/assays were PHE/NHSE approved. The reference antibody test was the Elecsys Anti-SARS-CoV-2 assay (Roche diagnostics GmBH).

**Main outcome measures:** Sensitivity and specificity of the rapid antibody test

**Results:** The reference antibody test was positive in 190/268 (70.9%) of participants with a history of symptoms suggestive of COVID-19; in the majority (n=312) the POC test was taken 35 days or more after onset of symptoms. The POC antibody test had an overall sensitivity of 90.1% (292/328, 95% CI 86.3 – 93.1) and specificity of 100% (68/68, 95% CI 94.7 - 100) for confirming prior SARS-CoV-2 infection when compared to the composite reference standard. Sensitivity was 97.8% (89/92, 95% CI 92.3% to 99.7%) in participants who had been admitted to hospital and 84.4% (124/147, 95% CI 77.5% to 89.8%) in those with milder illness who had never been seen in hospital.

**Conclusions:** The Livzon point-of-care antibody test had comparable sensitivity and specificity to the reference laboratory antibody test, so could be used in clinical settings to support decision-making about patients presenting with more than 10 days of symptoms of COVID-19.

**What is already known on this topic:**

- Presence of IgG and IgM antibodies to SARS-COV-2 indicates that the person was infected at least 7 days previously and is usually no longer infectious.
- Rapid point-of-care tests for antibodies to SARS-COV-2 are widely available, cheap and easy to use
- Preliminary evaluations suggested that rapid antibody tests may have insufficient accuracy to be useful for testing individual patients.

**What this study adds:**

- The rapid point-of-care test for antibodies to SARS-COV-2 was 90.1% sensitive and 100% specific compared to reference standards for prior infection with COVID-19.
- This is comparable to reference antibody tests
- The point-of-care test evaluated in this study could be used to support clinical decision-making in real time, for patients presenting with symptoms of possible COVID-19 with at least 10 days of symptoms.

## Introduction

The COVID-19 pandemic has infected over 37 million people globally, causing over 1 million deaths. The UK has one of the world’s highest death tolls, with over 600,000 cases and over 42,000 deaths, equating to 630 deaths per million population. The current “gold standard” for diagnosis is PCR on nasopharyngeal (or nose and throat) swabs, although sensitivity can be as low as 71% in the community^1^.

In May 2020, Public Health England approved two laboratory-based antibody tests and has since approved several more; however it often takes several days to obtain the results in clinical settings. Rapid point-of-care antibody (IgM and IgG) tests (using a drop of blood from a finger prick) have been developed and are being used and marketed in the UK. If accurate, they have the potential to accelerate assessment of prior exposure to COVID-19 in patients presenting to hospital or GPs (who often have already had symptoms for 7 or more days). They may be particularly useful in diagnosing later presentations (e.g. with chest infections post COVID, post-acute COVID), in screening health workers and other key workers before return to work, in screening household contacts of COVID-19 patients, in testing patients with atypical symptoms, and for screening in work places.

Preliminary evaluations suggested that rapid antibody tests may have insufficient sensitivity and specificity to detect antibodies to SARS-COV-2 in individual patient applications^2,3^. The Medicines and Healthcare products Regulatory Agency (MHRA) has raised concerns about false positives and told providers to stop selling point-of-care antibody tests for home testing^4^. Most of the published data is based on stored blood samples from hospitalised patients, with no indication of the duration of illness prior to sampling ^3,5,6^. It is to be expected that sensitivity of the tests will vary according to the patient population (outpatient versus inpatient; age group; patients on immunosuppressive treatments), duration of infection at the time of sampling, and the characteristics of the particular test in question.

This study aims to establish the real-world diagnostic accuracy of a rapid point-of-care antibody test compared to PCR and/or the Public Health England approved laboratory-based antibody tests, in patients with a history of symptoms consistent with COVID-19.

## Methods

### Study Design

This was a cross-sectional diagnostic test accuracy study in patients with history of known COVID-19 infection confirmed by laboratory RT-PCR (Reverse Transcriptase – Polymerase Chain Reaction) and hospital staff with a history of clinically-suspected COVID-19.

### Participants

Participants were recruited at Watford General Hospital (West Hertfordshire NHS Trust). The study included two groups. The first group consisted of patients who had either been admitted as inpatients or had been monitored remotely by the “virtual hospital”, and had confirmed COVID-19 on PCR (from 5^th^ March to 18^th^ June 2020). Eligible patients were approached consecutively by clinicians at the study site. This group was recruited retrospectively after the PCR test had been performed.

The second group consisted of consecutive volunteers from the hospital staff with history of clinically suspected COVID-19 (based on their reported symptoms), irrespective of whether they had had a PCR test, but who had (or were going to have) a standard venous antibody test (from 1^st^ May to 22^nd^ July 2020). This group was recruited prospectively.

In both cases we included only adults (age 18 and over), at least 7 days after onset of symptoms, and who gave written informed consent. There was some overlap between the groups as many participants had both a PCR and a venous antibody test.

Following informed consent, we extracted clinical data such as symptoms and date of onset of illness from the medical records of participating patients. Staff were asked to complete a short questionnaire about their demographic characteristics, past medical history, symptoms and severity of suspected COVID-19. Where the date of onset of symptoms could not be recalled precisely, it was estimated.

### Test Methods

The index test was the rapid point-of-care (POC) IgM / IgG colloidal gold lateral flow immunoassay (LFIA) manufactured by Livzon (Zhuhai, Guangdong, China). We selected this test because it was the only point of care test serological test available at the time and had been endorsed by a leading authority (the Chinese Federal Drug Authority) and the European Respiratory Society.

A drop of capillary blood was obtained from participants using a single-use disposable lancet to prick a fingertip. The blood was applied to the rapid antibody test, followed by the test solution. The test was only carried out by one of three members of staff who had received training in use of the test, to ensure a standard approach. It was read by two independent observers after 15 minutes. In case of disagreement between the observers the result was recorded as “inconclusive”.

We used a composite reference and evaluated the components of this individually. The first component was SARS-COV-2 RT-PCR. As all the participants in the first group had a positive PCR (which was the inclusion criterion), the researchers were aware of the reference test result when reading the rapid antibody test. Samples were tested on multiple RT-PCR platforms using the available molecular technology during the study time (PHE laboratories, GeneXpert^®^ system Xpert, Xpress SARS-CoV-2 and Source bioscience laboratory). All molecular platforms and assays were PHE/NHSE approved.

The second reference test was the Elecsys Anti-SARS-CoV-2 assay. This is an ECLIA (Electro Chemi Luminescent Immuno Assay) manufactured by Roche Diagnostics GmbH which uses a recombinant protein representing the nucleocapsid (N) protein of SARS-CoV-2. The assay is listed as CE marked and has been approved by Public Health England (PHE). Its overall sensitivity is 83.9% and specificity is 100%^7^. This was reported as being either positive or negative, without specifying IgM or IgG or titre. In most cases the reference test was done after the POC test; where this was not the case, the researcher and participant were usually unaware of the result of the reference test.

### Analysis

#### Sample size

For the first analysis, we aimed to recruit at least 138 patients with 7 or more days of symptoms and confirmed PCR-positive test for COVID-19. After day 7 of symptoms, 50% of patients are expected to have seroconverted; 90% are expected to have seroconverted by day 10 ^8^. With a predicted sensitivity of 90% after day 10, a minimum of 138 participants with symptom duration of at least 10 days were needed in order to be able to detect a sensitivity of 90% with a 95% confidence interval width of +/-5% ^9^. For the second analysis, we expected hospital staff undergoing a venous antibody test to have a prevalence of prior COVID-19 infection of around 30%. This analysis required a sample of at least 244 participants to determine sensitivity and specificity of at least 95% with a confidence interval of +/-5%. Therefore, we aimed to recruit at least 138 participants having had a positive PCR and 244 participants having had a reference antibody test.

#### Statistical analysis

The results of the rapid antibody test were cross tabulated against the reference standards. Sensitivity was calculated against each standard and against the composite reference standard and specificity was calculated against the reference antibody test and against the composite reference standard. We also calculated the area under the receiver-operator curve (AUROC), the positive predictive value (PPV) and negative predictive value (NPV). Inconclusive and missing results were excluded from these analyses.

The first analysis calculated the sensitivity of the POC test in participants with a positive PCR. The second analysis calculated sensitivity and specificity against the reference antibody test. Because antibodies may wane over time, we also conducted a sensitivity analysis for this, excluding cases where the interval between the POC test and the venous antibody test was greater than 28 days. For the third analysis, we calculated sensitivity and specificity of the POC text compared to the composite reference standard; “positive” was defined as a positive PCR test for COVID-19 before the rapid antibody test AND/OR a positive PHE - approved laboratory antibody test. The composite reference standard “negative” was defined as a negative PHE-approved antibody test, and the absence of any prior positive PCR test.

We conducted subgroup analyses in participants with presumed milder illness (who were never seen in hospital), compared to participants who had been admitted as inpatients. We also analysed subgroups in whom the POC test was conducted at least 20 days after onset of symptoms (the criterion set by the MHRA), and at least 35 days after onset of symptoms, a group in which published data is currently lacking^10^.

We also estimated the sensitivity of the reference standards by comparing them to each other. To evaluate the sensitivity of the PCR, we included participants with a positive venous antibody test who had also had a PCR test within 7 days of the onset of their symptoms. To estimate sensitivity of the venous antibody test, we included participants with a positive PCR who had also undergone a reference antibody test, at least 14 days after the onset of symptoms.

### Ethical approvals

This study was approved by the Faculty of Medicine Research Ethics Committee at the University of Southampton (reference 56480) and by the Wales Research Ethics Committee 4 (Wrexham, IRAS 283264, REC 20/WA/0148).

### Patient and public involvement

Health care workers who were patients themselves contributed to the design, funding and recruitment to this research. We plan to disseminate the results to study participants where it is possible to reach them.

## Results

### Participants

We recruited 398 participants in total (173 in the first group of patients and 225 in the second group of hospital staff). Two participants in the second group did not have a usable reference antibody test (one sample was misplaced and the other was taken only 4 days after the onset of symptoms) so were excluded. The characteristics of the remaining 396 participants are summarised in Table 1. About half of the participants (52.8%) had never been seen in hospital, whereas the others had been assessed in A&E, 20.6% were followed up in the “virtual hospital” and 23.6% were admitted as inpatients. Regarding the reference test, 130 participants had only a PCR test, 124 had only a reference antibody test, and 144 had both. Table 1 also shows the characteristics of the participants included in each analysis. Ninety-three participants were included in all of the analyses. The principal difference is that in the first group (with a positive PCR test) the majority of patients had been assessed or admitted to hospital; only 22.6% had never been assessed in hospital; whereas in the second group (with a venous antibody test) more than three-quarters (76.5%) had never been assessed in hospital. More of the second group were smokers and from BAME groups, but otherwise demographic characteristics were similar.

**Table 1:**
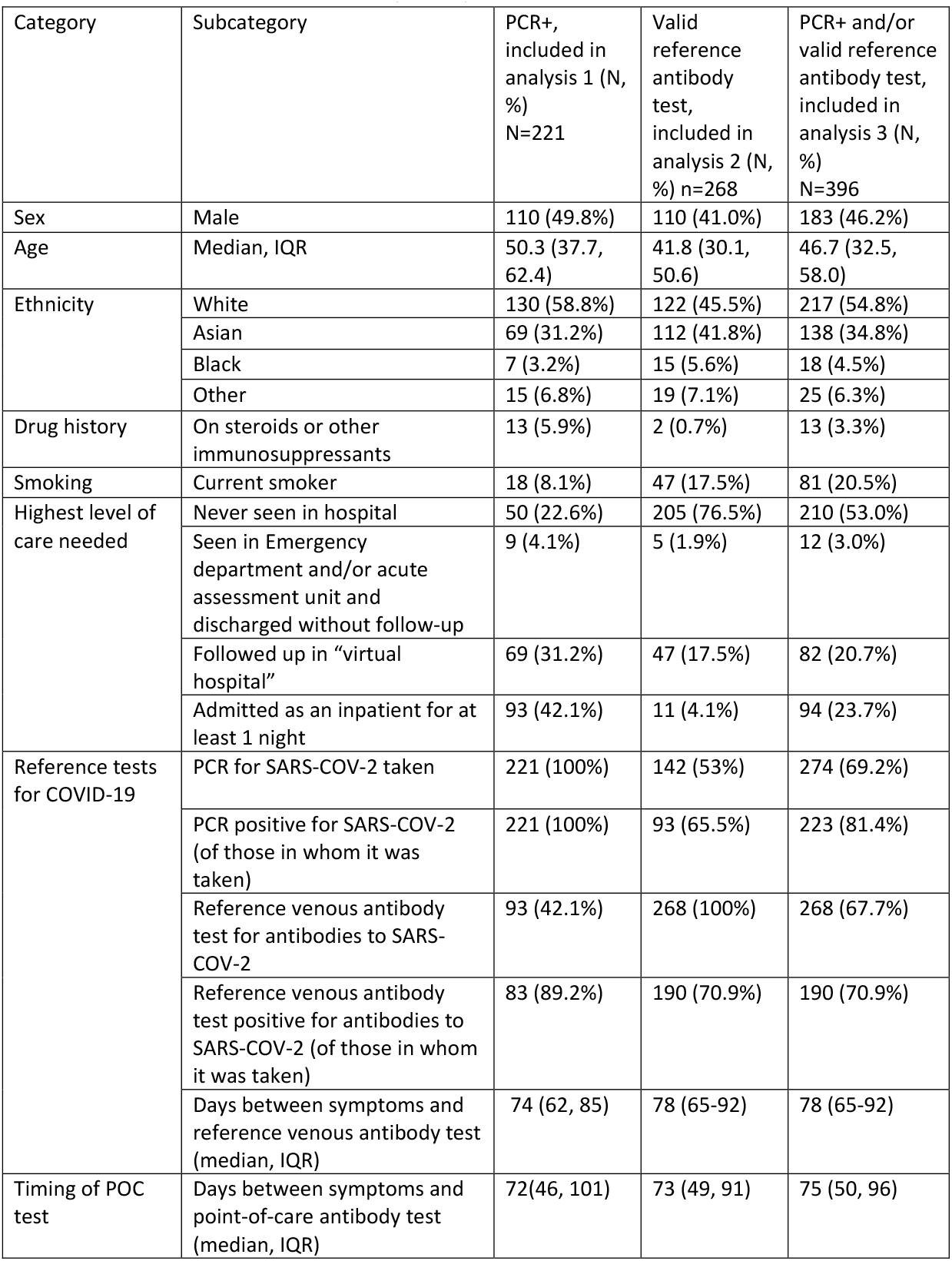
Baseline characteristics of participants

| Category | Subcategory | PCR+, included in analysis 1 (N, %) N=221 | Valid reference antibody test, included in analysis 2 (N, %) n=268 | PCR+ and/or valid reference antibody test, included in analysis 3 (N, %) N=396 |
| --- | --- | --- | --- | --- |
| Sex | Male | 110 (49.8%) | 110 (41.0%) | 183 (46.2%) |
| Age | Median, IQR | 50.3 (37.7, 62.4) | 41.8 (30.1, 50.6) | 46.7 (32.5, 58.0) |
| Ethnicity | White | 130 (58.8%) | 122 (45.5%) | 217 (54.8%) |
|  | Asian | 69 (31.2%) | 112 (41.8%) | 138 (34.8%) |
|  | Black | 7 (3.2%) | 15 (5.6%) | 18 (4.5%) |
|  | Other | 15 (6.8%) | 19 (7.1%) | 25 (6.3%) |
| Drug history | On steroids or other immunosuppressants | 13 (5.9%) | 2 (0.7%) | 13 (3.3%) |
| Smoking | Current smoker | 18 (8.1%) | 47 (17.5%) | 81 (20.5%) |
| Highest level of care needed | Never seen in hospital | 50 (22.6%) | 205 (76.5%) | 210 (53.0%) |
|  | Seen in Emergency department and/or acute assessment unit and discharged without follow-up | 9 (4.1%) | 5 (1.9%) | 12 (3.0%) |
|  | Followed up in “virtual hospital” | 69 (31.2%) | 47 (17.5%) | 82 (20.7%) |
|  | Admitted as an inpatient for at least 1 night | 93 (42.1%) | 11 (4.1%) | 94 (23.7%) |
| Reference tests for COVID-19 | PCR for SARS-COV-2 taken | 221 (100%) | 142 (53%) | 274 (69.2%) |
|  | PCR positive for SARS-COV-2 (of those in whom it was taken) | 221 (100%) | 93 (65.5%) | 223 (81.4%) |
|  | Reference venous antibody test for antibodies to SARS-COV-2 | 93 (42.1%) | 268 (100%) | 268 (67.7%) |
|  | Reference venous antibody test positive for antibodies to SARS-COV-2 (of those in whom it was taken) | 83 (89.2%) | 190 (70.9%) | 190 (70.9%) |
|  | Days between symptoms and reference venous antibody test (median, IQR) | 74 (62, 85) | 78 (65-92) | 78 (65-92) |
| Timing of POC test | Days between symptoms and point-of-care antibody test (median, IQR) | 72(46, 101) | 73 (49, 91) | 75 (50, 96) |

Among 274 participants who had a PCR test for SARS-COV-2, it was positive in 223 (81.4%), but two of these the PCR was taken after the POC test so were excluded from the analysis. The first analysis included only the 221 participants who had had a positive PCR test before the POC test. The median interval between the positive PCR test and the POC antibody test was 60 days (IQR 35-91).

Of 268 participants who had a reference venous blood test for antibodies to SARS-COV-2, the test was positive in 190 (70.9%). The median interval between the POC test and the reference antibody test was 3 days (IQR −10 to 24 days).

Most of the participants had the POC test at least 35 days after the onset of their symptoms. The date of onset of symptoms was missing for 45 participants (not recorded in hospital notes). For the remaining participants, the median interval between onset of symptoms and the POC antibody test was 74 days (IQR 50-96, range 7-173 days). Only 11 participants had the POC test taken 7-14 days after onset of symptoms (of whom only 2 <10 days), 11 had it taken at 11-21 days, 19 at 22-34 days, and the remaining 312 at 35 days or more.

### Accuracy of the point-of-care antibody test

Of the 221 participants who had a positive PCR test for COVID-19, the POC result was inconclusive in 5 for the IgG, 6 for the IgM and 3 overall. In the 218 for which there was a conclusive POC result, it was positive for IgG and/or IgM in 197 cases, corresponding to a sensitivity of 90.4% (95% CI: 85.7% to 93.9%, table 2).

**Table 2:**
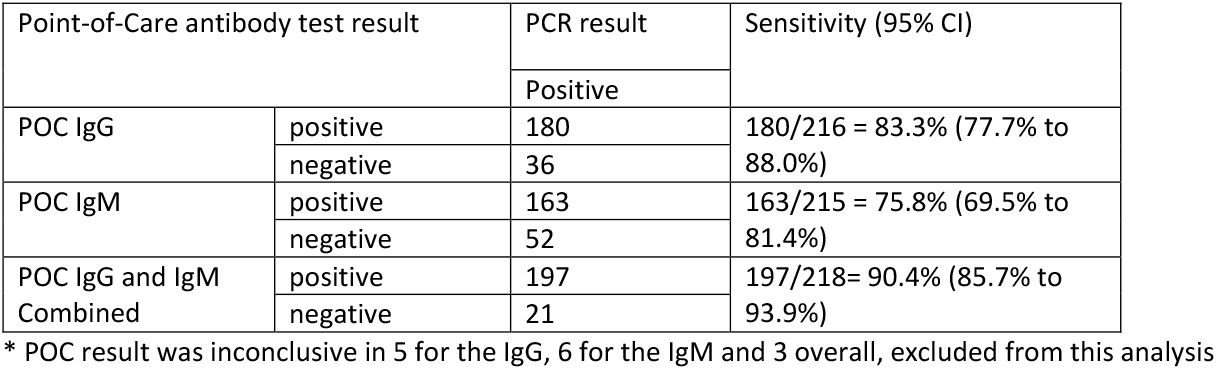
Analysis 1: Sensitivity of the POC antibody test in participants with positive PCR for SARS-COV-2 (n=221*)

Test accuracy relative to the reference venous antibody test is shown in table 3. Using the combined IgG and/or IgM, the POC test had a sensitivity of 92.0% (95% CI 87.2 – 95.5) and specificity of 98.7% (95% CI 93.1 to 100). Our sensitivity analysis, excluding cases where the interval between the POC test and the venous antibody test was greater than 28 days, resulted in a sensitivity of 137 / 148 = 92.6% (95%CI 87.1% to 96.2%) and specificity of 58/59 = 98.3% (95% CI 90.9% to 100.0%).

**Table 3:** Analysis 2: Accuracy of the POC antibody test compared to the reference laboratory antibody test (n=268*)

| Point-of-Care antibody test result |  | Reference Antibody result |  | Sensitivity (95% CI) | Specificity (95% CI) | ROC (95% CI) | PPV (95% CI) | NPV (95% CI) |
| --- | --- | --- | --- | --- | --- | --- | --- | --- |
|  |  | Positive | Negative |  |  |  |  |  |
| POC IgG | positive | 147 | 0 | 147/187 = 78.6% (72.0% to 84.3%) | 78/78 = 100% (95.4% to 100%) | 0.89 (0.86, 0.92) | 100% (97.5% to 100%) | 66.1% (56.8% to 74.6%) |
|  | negative | 40 | 78 |  |  |  |  |  |
| POC IgM | positive | 139 | 1 | 139/184 = 75.5% (68.7% to 81.6%) | 77/78 = 98.7% (93.1% to 100%) | 0.87 (0.84, 0.90) | 99.3% (96.1% to 100%) | 63.1% (53.9% to 71.7%) |
|  | negative | 45 | 77 |  |  |  |  |  |
| POC IgG and IgM Combined | positive | 173 | 1 | 173/188 = 92.0% (87.2% to 95.5%) | 77/78 = 98.7% (93.1% to 100%) | 0.95 (0.93, 0.98) | 99.4% (96.8% to 100%) | 83.7% (74.5% to 90.6%) |
|  | negative | 15 | 77 |  |  |  |  |  |

\* POC result was inconclusive in 3 for the IgG, 6 for the IgM and 2 overall

POC test accuracy relative to the composite reference standard is given in table 4. The POC had an overall sensitivity of 90.1% (292/328, 95% CI 86.3 – 93.1) and specificity of 100% (68/68, 95% CI: 94.7% to 100%). In the subgroup of participants with presumed milder illness (who were never seen in hospital), the sensitivity was 84.4% (124/147, 95% CI 77.5% to 89.8%) whereas in patients admitted to hospital, the sensitivity was 97.8% (89/92, 95% CI 92.3% to 99.7%). In the subgroup where the POC test was conducted at least 20 days after onset of symptoms, sensitivity was 91.1% (288/316, 95% CI 87.4% to 94.0%). This was very similar (90.8%, 277/305, 95% CI 87.0% to 93.8%,) in the subgroup where the POC test was conducted at least 35 days after onset of symptoms. Numbers were too small to calculate diagnostic accuracy in the subgroups of patients in whom the POC test was taken at 7-14 days, 15-21 and 22-34 days after onset of symptoms.

**Table 4:**
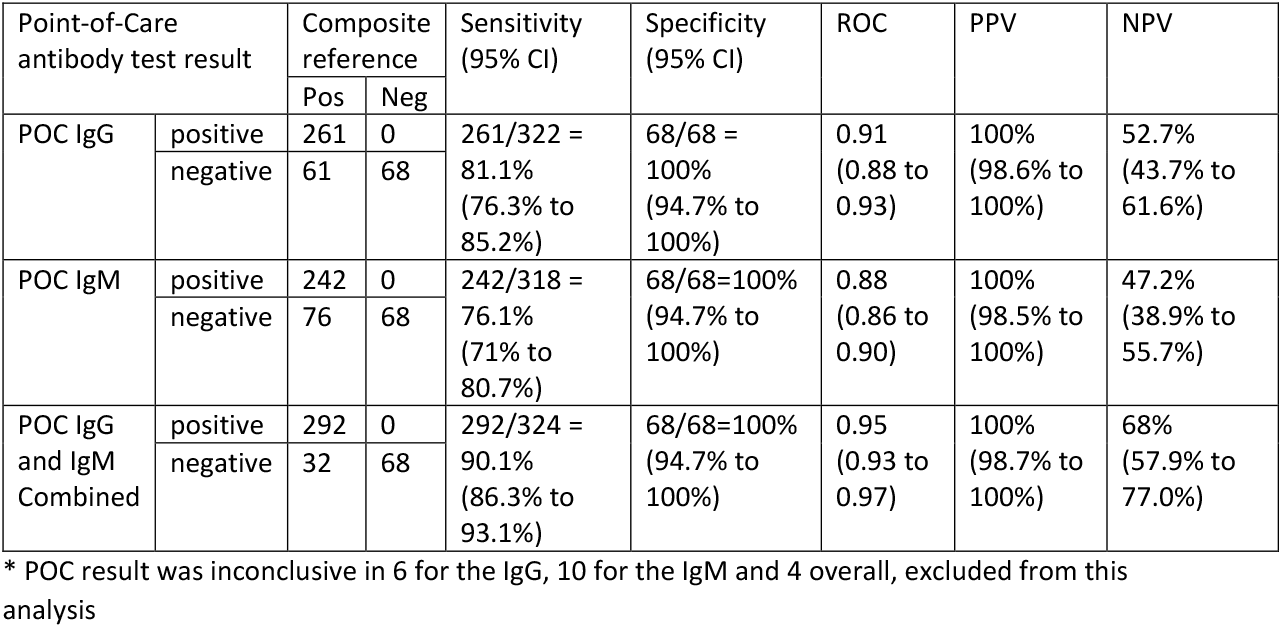
Accuracy of the POC antibody test compared to the composite reference standard (PCR and reference antibody, n=396*)

| Point-of-Care antibody test result |  | Composite reference |  | Sensitivity (95% CI) | Specificity (95% CI) | ROC | PPV | NPV |
| --- | --- | --- | --- | --- | --- | --- | --- | --- |
|  |  | Pos | Neg |  |  |  |  |  |
| POC IgG | positive | 261 | 0 | 261/322 = 81.1%<br>(76.3% to 85.2%) | 68/68 = 100%<br>(94.7% to 100%) | 0.91<br>(0.88 to 0.93) | 100%<br>(98.6% to 100%) | 52.7%<br>(43.7% to 61.6%) |
|  | negative | 61 | 68 |  |  |  |  |  |
| POC IgM | positive | 242 | 0 | 242/318 = 76.1%<br>(71% to 80.7%) | 68/68=100%<br>(94.7% to 100%) | 0.88<br>(0.86 to 0.90) | 100%<br>(98.5% to 100%) | 47.2%<br>(38.9% to 55.7%) |
|  | negative | 76 | 68 |  |  |  |  |  |
| POC IgG and IgM Combined | positive | 292 | 0 | 292/324 = 90.1%<br>(86.3% to 93.1%) | 68/68=100%<br>(94.7% to 100%) | 0.95<br>(0.93 to 0.97) | 100%<br>(98.7% to 100%) | 68%<br>(57.9% to 77.0%) |
|  | negative | 32 | 68 |  |  |  |  |  |

\* POC result was inconclusive in 6 for the IgG, 10 for the IgM and 4 overall, excluded from this analysis

There were no adverse events from performing the rapid POC tests or the reference standards. The test was easy to use in a routine clinical setting and gave results within 15 minutes, whereas it took several days to receive results of the reference antibody test.

### Sensitivity of the reference standards

Of 54 patients with a positive venous antibody test who also had a PCR within 7 days of onset of symptoms, 45 had a positive PCR. This implies that the sensitivity of the PCR test was 83.3% (95% CI 70.7 – 92.1) in this cohort. Of 93 patients with a positive PCR who had a subsequent venous antibody test (at least 14 days after onset of symptoms), 83 had a positive reference antibody test (sensitivity of 89.2%, 95% CI 81.1 – 94.7).

## Discussion

### Principal findings

The reference antibody test was positive in 70.9% of participants with a history of symptoms of COVID-19. The Livzon POC antibody test was highly specific for confirming prior COVID-19 infection when compared to both PCR and laboratory-based antibody testing, with no false positive results. Sensitivity compared to the composite reference test was 90.1%. Sensitivity was higher in participants who had been admitted to hospital than in those who were never seen in hospital.

### Strengths and weaknesses of the study

This study was conducted in a true “point-of-care” setting, not using laboratory samples. This shows that the test had good diagnostic test accuracy when used in a routine clinical setting. We included a good balance of patients, both admitted to hospital and never seen in hospital, which also makes the findings more widely generalisable. All participants had clinical symptoms of COVID-19, and there is information about the course of their illness which enables us to examine diagnostic accuracy in different subgroups. However, while it is possible to provide a “gold standard” in laboratory settings using stored sera, this is not possible in a clinical setting as there is no “gold standard” which is 100% sensitive and specific in clinical practice. According to official documentation, the overall sensitivity of the reference venous antibody test is 83.9% overall, 86.1% at or after 14 days, or 86.7% at or after 21 days of symptom onset^7^. Our own data confirms that compared to PCR, the reference antibody test was 89.2% sensitive. Although a positive PCR can be used as a gold standard to determine sensitivity, the PCR itself is not 100% sensitive and cannot be used as a “gold standard” for determining specificity^11^. However, a strength of our study is that many of our participants had both a PCR test and a venous antibody test; using these in combination improves the accuracy of the reference standard.

As the POC test was conducted by clinicians looking after the participants, it was not possible to blind them to the result of the PCR, in particular in the first group where a positive PCR for COVID-19 was an inclusion criterion. This decision was taken at the start of the study because at the time there was no approved reference antibody test, and the main aim for the first group was to determine sensitivity.

As this was a “real-word” evaluation, some data were missing. In some cases there was a lengthy interval between symptoms and the point-of-care test, and between the point-of-care test and the reference venous antibody test. This was because the reference antibody test was only available from May 2020 and some participants first experienced symptoms in February and early March 2020. However, a sensitivity analysis excluding cases where the interval between the POC test and venous antibody test was >28 days still gives a similar sensitivity.

Another limitation is that our cohort was relatively young (median age was 46.7); almost all of our participants aged 65 or over were inpatients with a positive PCR. It is uncertain whether comparable results would be seen in elderly patients in the community, who may not manifest a good antibody response. The tests were only evaluated in one centre and from one batch. There have been reports of variation in sensitivity between batches of rapid antibody tests^12^ so good quality control will be essential.

### Strengths and weaknesses in relation to other studies, discussing important differences in results

A recent systematic review concluded that “available evidence does not support the continued use of existing point-of-care serological tests”, because their overall sensitivity was found to be low at 66% (95% CI 49.3 – 79.3)^3^. However, most of the studies were small and under-powered. Only four of the LFIA studies included in this review included outpatients; the total number of participants in these was the same as the number of participants in our study (398). Many of the included studies used samples taken from patients early in the course of their infection, when antibodies are not expected to have developed in all patients; this may partly explain the lower reported sensitivity. This study has a larger sample size than many of the earlier publications, and a larger number of COVID-19 patients, not only inpatients but also many participants who were never admitted to hospital.

A recent study evaluated 10 commercially available POC tests and showed variable reported sensitivities ranging from 81.0%-95.2% at 14 or more days after symptom onset and specificity ranging from 75.7%-99.2%^13^; only a third of the tests met the acceptable sensitivity and specificity thresholds. In our study, the Livzon POC test was very sensitive with no false positives. There are several previous evaluations of the same Livzon POC test. The first evaluation in China included 86 positive and 22 negative patients (all inpatients), and the tests were all conducted in the laboratory; sensitivity was 92.9% (95% CI 75.0 – 98.0) at 7-14 days and 96.8% (95% CI 81.5 – 99.8) at 15 days or more after onset of symptoms^14^. An assessment for the US Federal Drug Administration (FDA) found a sensitivity of 86.7% (95% CI 70.3 – 94.7%) and specificity of 97.5% (95% CI 91.3 – 99.3%) using 30 positive and 80 negative stored sera in a laboratory (two false positive IgM results were reported)^15^. An evaluation in French inpatients (1-25 days after onset of illness) found a sensitivity of 86.7% (95% CI 69.5 – 100) and specificity of 100% for the Livzon rapid IgG (but one false positive IgM was reported)^16^. In the current study, we provide much more data on the Livzon point-of-care antibody test, especially for patients over 35 days after onset of symptoms, confirming that the rapid antibody tests remain sensitive at this time.

### Meaning of the study: possible explanations and implications for clinicians and policymakers

Point-of-care COVID-19 antibody tests for home testing were withdrawn from sale in the UK at the end of May 2020 because it was felt that the laboratory tests have a “much higher standard of accuracy” ^4^. There may be issues with processing of samples and results by providers of “home testing kits” which our study was unable to evaluate, but it seems that in the hands of clinicians, the Livzon point-of-care test has a similar level of accuracy to the approved laboratory tests. The role of IgM in diagnosing early infection is still not fully elucidated as it does not consistently appear before IgG. However, the POC tests are cheaper, quicker, and are a useful addition to the diagnostics available to clinicians on the frontline.

For patients with onset of symptoms at least 10 days previously, the POC antibody test can provide rapid and reliable information as to whether a patient has previously had COVID-19. This can be particularly useful in settings where access to PCR is limited and patients are unable to access a PCR within their first 10 days of illness. Given the problems with PCR sensitivity, especially if conducted after the first week of illness, the Livzon antibody test may also have a role in diagnosing people who have had symptoms for 10 days or more, whether or not they have had a PCR test. In a patient with a high index of suspicion (and symptoms for 10 days or more) but a negative PCR, an antibody test may be of more value than repeating the PCR. The combination of PCR plus IgG and IgM testing in suitable patients has already been suggested to improve diagnostic accuracy^17^.

In primary care, the test would help to inform decisions about clinical management and return to work in patients with ongoing symptoms of “post-acute” COVID-19^18^. Being able to reassure the patient that they are still exhibiting investigation findings consistent with recent infection will be reassuring if the rest of the assessment is satisfactory.

In an occupational health setting, the use of POC testing in large workplaces such as factories and food processing plants may help to support return to work of post symptomatic patients. By day 10 in a recovered patient, the probability of positive viral culture is low^19^ (hence low infectivity) and furthermore when neutralising antibodies are detected (at a titre of 1 in 80 or higher), patients are no longer shedding infectious virus^20^. Thus, by confirming both an IgG and IgM response in a recovered individual, the time point of the patient’s disease can be more closely estimated as being above 10 days and it is very unlikely that the patient is still infectious. However, as clinical trials of vaccines against SARS-COV-2 recruit many more participants, it will be important to enquire about history of vaccination, as this will also result in an IgG response^21^.

### Unanswered questions and future research

Our understanding of post COVID-19 immunity is limited and constantly evolving-given increasing evidence of fading immune responses as seen in other coronaviruses, it would appear unlikely that patients with a positive response could be reassured or assume they have long lasting immunity. In patients infected with SARS-CoV-1, 90% have IgG for up to 2 years and 50% for up to 3 years^22^. It is unknown whether the antibodies detected by the POC test (or any antibody tests currently available) provide protection or immunity against COVID-19, or how long the antibodies will persist. Further research is needed to determine this. Nevertheless, an immediate priority would be to confirm whether patients with a recent history of symptoms and a positive IgM/IgG antibody test to SARS-COV-2 are no longer infectious and can therefore be advised they no longer need to self-isolate. If so, this could be an important tool in helping to manage the current pandemic.

**Figure 1:**
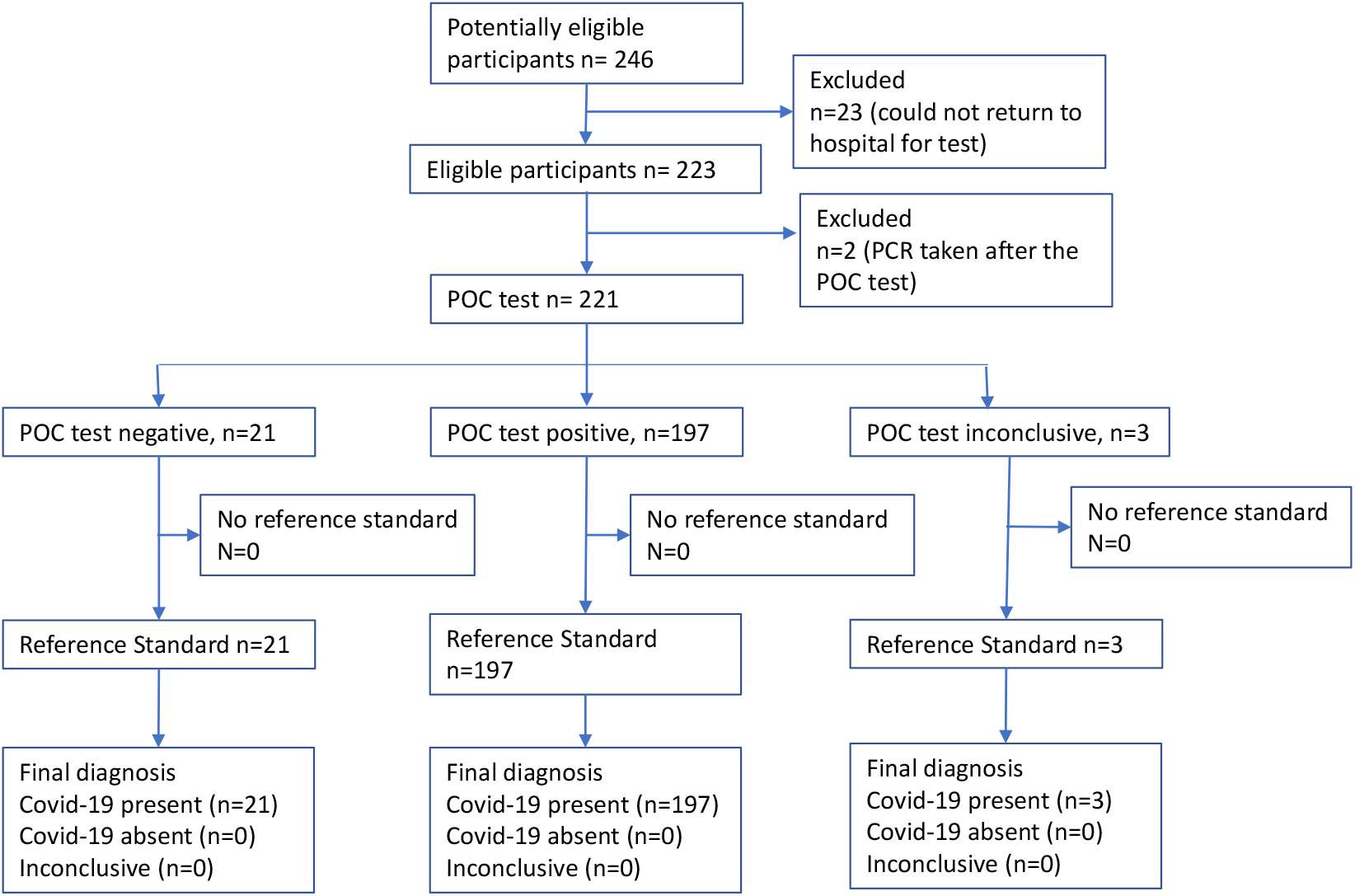
Flowchart for analysis 1 (participants with positive PCR for COVID-19)

**Figure 2:**
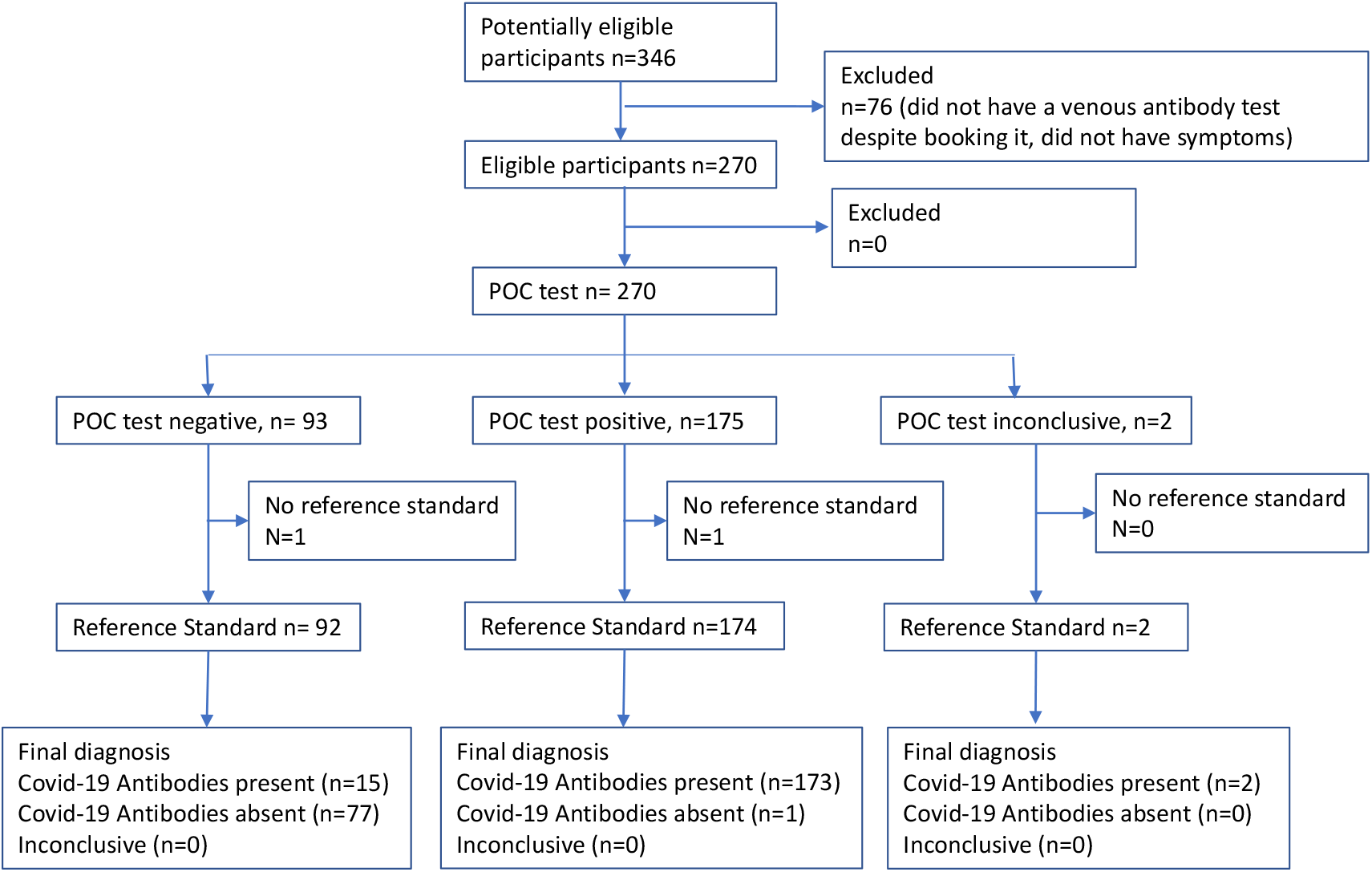
Flowchart for analysis 2 (participants who had a reference antibody test for COVID-19)

**Figure 3:**
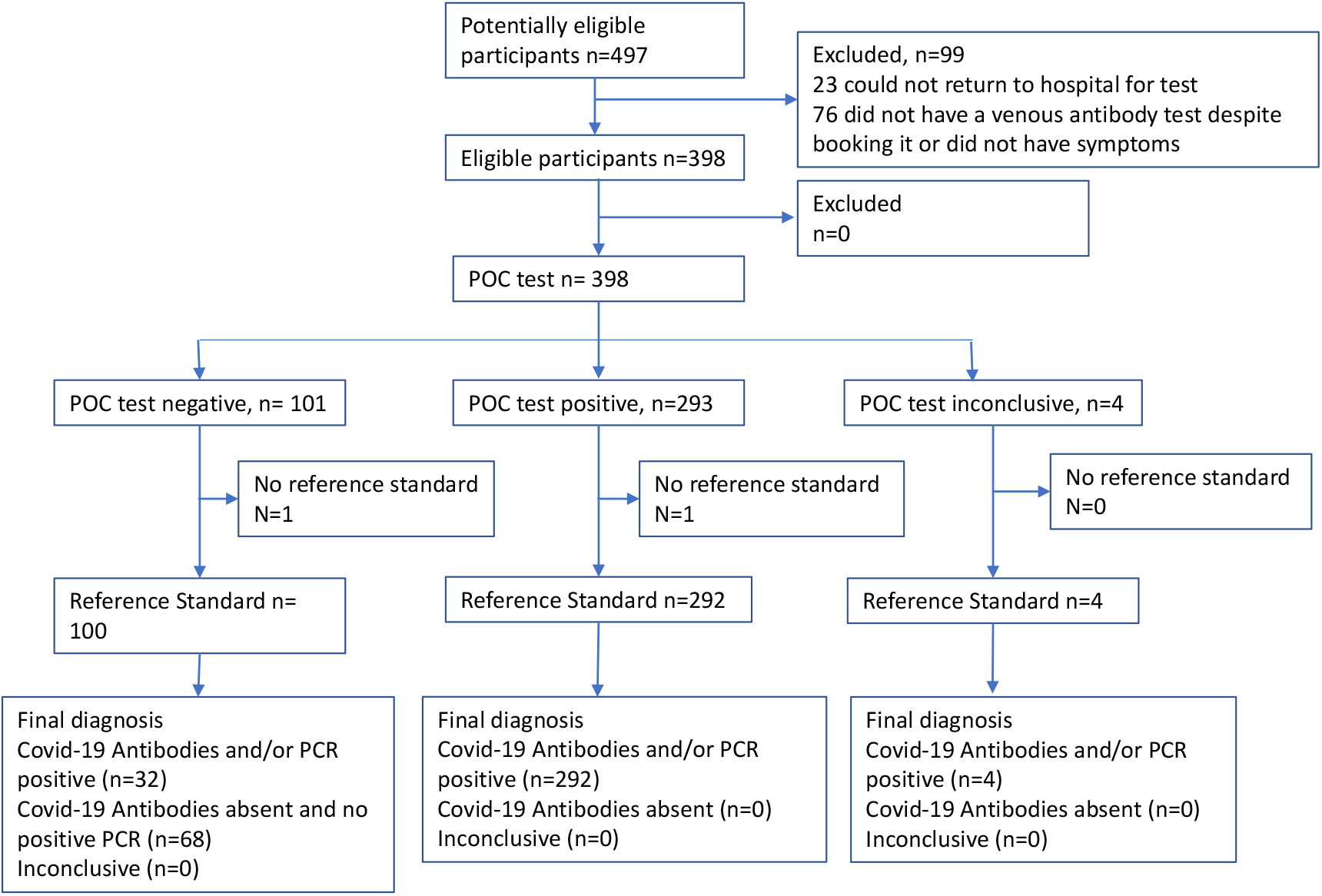
Flowchart for analysis 3 (composite reference standard)

## Conclusion

A rapid fingerprick point-of-care test for antibodies to SARS-COV-2 was 90.1% sensitive and 100% specific compared to the composite reference standard of PCR and reference antibody test on venous blood. Sensitivity was 97.8% in patients with a history of admission to hospital and 84.4% in those never assessed in hospital. This test may have a role in assisting primary care practitioners managing post-acute/long COVID, occupational health settings and in the acute setting, especially where access to PCR is limited.

## Supporting information

STARD checklist

## Data Availability

Data is available from the corresponding author on reasonable request.

## Acknowledgements

We would like to thank Wendy O’Brien and Clare McDermott at the School of Primary Care and Catherine Simpson at Southampton Clinical Trials Unit at the University of Southampton who supported the trial management for this study. Rosie Bourke and Alex Newland Smith arranged follow up for the first group to have the POC tests. The POC tests were funded by The Medical Director’s cardiology charity and by individuals donating to a “Just Giving” page. We received no commercial funding or free tests from the manufacturer. West Hertfordshire Hospitals NHS Trust supported clinical staff time. MLW’s salary was funded by the National Institute of Health Research (NIHR), under grant CL-2016-26-005. Southampton CTU staff were supported by NIHR CTU Support Funding. The funders had no role in this research.

